# Antiviral efficacy of fluoxetine in early symptomatic COVID-19: an open-label, randomised, controlled, adaptive platform trial (PLATCOV)

**DOI:** 10.1101/2024.01.16.24301337

**Authors:** Podjanee Jittamala, Simon Boyd, William HK Schilling, James A Watson, Thundon Ngamprasertchai, Tanaya Siripoon, Viravarn Luvira, Elizabeth M Batty, Phrutsamon Wongnak, Lisia M Esper, Pedro J Almeida, Cintia Cruz, Fernando R Ascencao, Renato S Aguiar, Najia K Ghanchi, James J Callery, Shivani Singh, Varaporn Kruabkontho, Thatsanun Ngernseng, Jaruwan Tubprasert, Wanassanan Madmanee, Kanokon Suwannasin, Amornrat Promsongsil, Borimas Hanboonkunupakarn, Kittiyod Poovorawan, Manus Potaporn, Attasit Srisubat, Bootsakorn Loharjun, Walter RJ Taylor, Farah Qamar, Abdul Momin Kazi, M. Asim Beg, Danoy Chommanam, Sisouphanh Vidhamaly, Kesinee Chotivanich, Mallika Imwong, Sasithon Pukrittayakamee, Arjen M Dondorp, Nicholas PJ Day, Mauro M Teixeira, Watcharapong Piyaphanee, Weerapong Phumratanaprapin, Nicholas J White, the PLATCOV Collaborative Group

## Abstract

**Background:** The selective serotonin reuptake inhibitors (SSRIs) fluoxetine and fluvoxamine were repurposed for the treatment of early COVID-19 based on their antiviral activity *in vitro*, and observational and clinical trial evidence suggesting they prevented progression to severe disease. However, these SSRIs have not been recommended in guidelines and their antiviral activity *in vivo* has not been characterised.

**Methods:** PLATCOV is an open-label, multicentre, phase 2, randomised, controlled, adaptive pharmacometric platform trial running in Thailand, Brazil, Pakistan, and Laos. We recruited low-risk adult outpatients aged 18-50 with early symptomatic COVID-19 (symptoms <4 days). Patients were assigned using block randomisation to one of eleven treatment arms including oral fluoxetine (40mg/day for 7 days), or no study drug. Uniform randomisation ratios were applied across the active treatment groups while the no study drug group comprised ≥20% of patients at all times.

The primary endpoint was the rate of oropharyngeal viral clearance assessed in a modified intention-to-treat population (>2 days follow-up). The viral clearance rate was estimated under a Bayesian hierarchical linear model fitted to the log10 viral densities in standardised duplicate oropharyngeal swab eluates taken daily over one week (18 measurements per patient). This ongoing trial is registered at ClinicalTrials.gov (NCT05041907).

**Findings:** Between 5 April 2022 and 8 May 2023 271 patients were concurrently randomised to either fluoxetine (n=120) or no study drug (n=151). Fluoxetine was well tolerated and accelerated the rate of viral clearance relative to the no study drug arm by 15% (95% credible interval (CrI): 2% to 34%). In a pooled meta-analysis including all unblinded patients the antiviral activity of fluoxetine was substantially less than ritonavir-boosted nirmatrelvir-85% increase in rate of viral clearance (95% CrI: 61 to 112%); and less than remdesivir 35% (14 to 59%), molnupiravir 37% (18 to 60%), and casirivimab/imdevimab 29% (10 to 48%).

**Interpretation:** Fluoxetine has *in vivo* antiviral activity against SARS-CoV-2. Although the level of antiviral efficacy is substantially less than with other currently available antiviral drugs, fluoxetine might still be useful in prophylaxis where less antiviral effect is required.

**Funding:** Wellcome Trust Grant ref: 223195/Z/21/Z through the COVID-19 Therapeutics Accelerator.

**Evidence before this study:** The SSRIs fluoxetine and fluvoxamine have been proposed as COVID-19 therapeutics based initially on observational, randomised trial and *in vitro* evidence. The observational reports suggested that patients taking SSRIs had a reduced probability of developing severe COVID-19 and dying. We searched PubMed and EMBASE for studies in English up until the 30^th^ November 2023 using the search terms “fluoxetine”, “fluvoxamine” and “COVID-19” with the search restricted to randomised controlled trials (RCTs). Eight outpatient fluvoxamine RCTs were identified. There were no fluoxetine RCTs in outpatients. A meta-analysis of available RCTs is compatible with a moderate reduction in hospitalisation and death in COVID-19 patients with an estimated risk ratio of 0.80 (95% CI: 0.62,1.01).

**Added value of the study:** We showed that in early COVID-19 illness the SSRI fluoxetine has weak antiviral activity *in vivo*. This activity is substantially less than other available antivirals such as ritonavir-boosted nirmatrelvir and molnupiravir. The pharmacometric approach described here provides a quantitative measure of *in vivo* antiviral effects with tractable sample sizes.

**Implications of available evidence:** Fluoxetine has weak *in vivo* antiviral activity in early COVID-19. This is insufficient for treatment but, as less antiviral activity is required to prevent an infection, fluoxetine could still be beneficial in prophylaxis.

## Introduction

Repurposing of existing small molecule drugs can provide affordable and widely available treatment options. At the beginning of the COVID-19 pandemic there was considerable interest in drug repurposing, but there was little success in demonstrating clinical efficacy apart from the use of immunomodulatory drugs for severe and hospitalised patients (e.g. dexamethasone),^1^ No clear benefits were demonstrated for the initial antiviral candidates. Now, four years later, there are several approved efficacious antiviral drugs to treat early symptomatic COVID-19, but these are expensive, and they are not widely available. Ritonavir-boosted nirmatrelvir is currently the most effective small molecule antiviral drug but, apart from its very high cost (up to $500 USD/course), it has other drawbacks including drug interactions, dysgeusia, and is associated with viral rebound. The only other widely available efficacious oral drug, molnupiravir, has concerns over generation of mutant viruses.^2,3^ There remains a need for effective, reliable, accessible, and affordable antiviral treatments for early COVID-19.

Selective serotonin reuptake inhibitors (SSRIs) are the most widely used class of antidepressants. They are readily available and affordable globally. In some countries over 10% of the adult population are taking SSRIs. Observational studies early in the pandemic suggested that patients taking fluoxetine had reduced mortality when admitted to hospital with COVID-19.^4,5^ Subsequent studies supported this observation,^6,7^ and also suggested that SSRIs confer a prophylactic benefit.^8^ Another SSRI, fluvoxamine, was assessed in a meta-analysis of 8 randomised clinical trials,^9–16^ (there were no outpatient randomised controlled trials for fluoxetine; supplementary appendix subsection S12). Treatment with fluvoxamine was compatible with a moderate reduction in hospitalisation or death in COVID-19 outpatients, with an estimated risk-ratio of 0.80 (95% CI: 0.62 to 1.01, Figure S2).

The proposed mechanism of action of SSRIs is through functional inhibition of acid sphingomyelinase (so-called FIASMAs). This interferes with intracellular endolyosomal viral trafficking.^17^Although many drugs are classified as FIASMAs, and some of the earlier research focussed on the closely related compound fluvoxamine, fluoxetine, which is on the WHO’s list of Essential Medications,^18^ was found to have the greatest *in vitro* FIASMA activity, the best tolerability profile, and the most favourable pharmacokinetic properties.^19^ *In vitro* anti-SARS-CoV-2 activity has been shown at fluoxetine concentrations approximating those during the treatment of depression (20 mg daily; 0.8 µg/ml, 2.6 µM).^20^ Pharmacokinetic modelling determined that an adult dose of 40 mg per day would provide at least 85% of patients with the trough target plasma concentrations needed to reach the estimated target 90% maximal effective concentration (EC90) within 3 days,^21^ although the justification for the extrapolated concentration target is not strong.

It is no longer feasible to conduct randomised trials assessing prevention of hospitalisation and death in outpatients with symptomatic COVID-19, as was done earlier in the pandemic. Even in high-risk patients, the proportion of patients with COVID-19 who progress to severe illness and/or require hospitalisation is now very low (<1%).^22^ For drugs with weak or moderate antiviral activity (such as fluoxetine) the sample sizes needed to show a clinical benefit have therefore become prohibitively large. An alternative approach for candidate antiviral drugs in early COVID-19 is to assess their *in vivo* pharmacodynamics i.e. their effects on the rate of viral clearance. Acceleration in viral clearance correlates with clinical benefit.^23,24^

PLATCOV is an adaptive platform trial in adults with acute early COVID-19. The PLATCOV trial methodology can evaluate antiviral activity rapidly and compare available treatments quantitatively.^2,25–27^ Here we report the results for fluoxetine and contextualise this by pooling all unblinded data from the platform and comparing fluoxetine with the other assessed antiviral interventions.

## Methods

### Study design

PLATCOV is an ongoing phase 2, open label, multicentre, randomised, controlled, adaptive platform trial running currently in Thailand, Brazil, Pakistan, and Laos (ClinicalTrials.gov: NCT05041907). The trial provides a standardised quantitative comparative methodology for *in vivo* assessment of potential antiviral treatments in low-risk adults with early symptomatic COVID-19. Potential antiviral treatments are entered into the platform when they become available, and they are removed when the prespecified stopping rules are reached. Enrolled patients were admitted to the study ward or managed as outpatients according to patient preference (none of the admissions were for clinical reasons, but for ease of adherence with the study procedures, or for self-isolation).

Standard symptomatic treatment was provided to all patients. Initially, the following drugs were studied: ivermectin, favipiravir, remdesivir, and casirivimab/imdevimab (monoclonal antibody cocktail). These groups have already reached the prespecified stopping rules for efficacy or lack of efficacy and so have been stopped^2,25–27^. Additional groups; ensitrelvir, molnupiravir, ritonavir-boosted nirmatrelvir, and the tixagevimab/cilgavimab monoclonal antibody cocktail, were introduced later. The primary analysis reported here includes the results from patients who were allocated concurrently to fluoxetine or no study drug (negative control). In addition, we present a meta-analysis of all small molecule drugs and monoclonal antibodies with unblinded data to provide a calibration of the effect sizes observed for fluoxetine.

PLATCOV is coordinated and monitored by the Mahidol Oxford Tropical Medicine Research Unit (MORU) in Bangkok, is overseen by a trial steering committee (TSC), conducted according to Good Clinical Practice principles, and approved by the local IRB/EC (see supplementary materials subsection S2). The results were reviewed regularly by a data and safety monitoring board (DSMB). The funders had no role in the design, conduct, analysis, or interpretation of the trial.

### Participants

Previously healthy non-pregnant adults aged between 18 and 50 years were eligible for enrolment in the trial if they had early symptomatic COVID-19 (i.e. symptoms for <4 days), oxygen saturation ≥96%, were unimpeded in activities of daily living, and gave fully informed written consent. SARS-CoV-2 positivity was defined either as a nasal lateral flow antigen test which became positive within two minutes (STANDARD® Q COVID-19 Ag Test, SD Biosensor, Suwon-si, Korea) or a positive PCR test with a cycle threshold value (Ct) <25 (all viral gene targets) within the previous 24 hours. Both tests ensure the majority of recruited patients have high viral loads. Exclusion criteria included taking any potential antivirals or pre-existing concomitant medications, chronic illness or significant comorbidity, haematological or biochemical abnormalities (haemoglobin <8 g/dL, platelet count <50,000/μL, abnormal liver function tests, and estimated glomerular filtration rate <70 mL/min per 1.73 m^2^), pregnancy (a urinary pregnancy test was performed in females), breastfeeding, or contraindication or known hypersensitivity to any of the study drugs.

### Randomisation and interventions

Block randomisation was performed via a centralised web-app designed by MORU software engineers using RShiny® hosted on a MORU webserver (supplementary appendix subsection S8). At enrolment, after obtaining fully informed consent and entering the patient details, the app provided the study drug allocation. The “no study drug” arm was allocated to a minimum proportion of 20% of patients, with uniform randomisation ratios applied across the other active treatment arms. The trial was open label as it was impractical to conceal the different interventions. The viral densities were measured blinded to treatment allocation. Fluoxetine was added to the platform on the 5^th^ May 2022 in Thailand, and the 21^st^ June 2022 in Brazil, Laos, and Pakistan. Fluoxetine was removed on the 8^th^ May 2023. During this period, patients were also randomised to remdesivir (until 10^th^ June 2022), casirivimab/imdevimab (Thailand only, until 20^th^ October 2022), favipiravir (until 30^th^ October 2022), molnupiravir (until 22^nd^ February 2023), tixagevimab/cilgavimab (until 4^th^ July 2023), nitazoxanide (Brazil, Laos and Pakistan, from 18^th^ January 2022 ongoing), ensitrelvir (Thailand and Laos only, from 17^th^ March 2023, ongoing), and ritonavir-boosted nirmatrelvir (from 6^th^ June 2022, ongoing as positive control).

### Procedures

All study drugs were stored under the appropriate conditions. Fluoxetine (Anzac ®: Bangkok Lab Cosmetic in Thailand and Laos, Prozac ®: Eli Lilly in Brazil and Flux, Hilton Pharma in Pakistan) was given at an oral dose of 40 mg per day for a total of seven days starting at baseline. The administration of all drugs was observed directly or via video. After randomisation and baseline procedures (see appendix page 9) oropharyngeal swabs (two swabs taken from each tonsil) were taken as follows. A flocked swab (Thermo Fisher MicroTest [Thermo Fisher, Waltham, MA, USA] and later COPAN FLOQSwabs® [COPAN Diagnostics, Murrieta, CA, USA]), was rotated against the tonsil through 360° four times and placed in Thermo Fisher M4RT (Thermo Fisher, Waltham, MA, USA) viral transport medium (3 mL). The swabs were transferred at 4–8°C, aliquoted, and finally frozen at –80°C within 48 hours. Separate swabs from each tonsil were taken once daily from day 0 to day 7, on day 10, and on day 14. Swabs were processed and tested separately. Vital signs were recorded three times daily by the patient (on the first day the initial vital signs were recorded by the study team). Symptoms and any adverse effects were recorded daily.

The TaqCheck® SARS-CoV-2 Fast PCR Assay (Applied Biosystems, Thermo Fisher Scientific, Waltham, MA, USA) quantitated viral loads (RNA copies per mL). This multiplexed real-time PCR method detects the SARS-CoV-2 N and S genes, and human RNase P gene in a single reaction. RNase P was used to adjust for variation in sample human cell content (see appendix page 20). Viral loads were quantified against ATCC (Manassas, VA, USA) heat-inactivated SARS-CoV-2 (VR-1986HK strain 2019-nCoV/USA-WA1/2020) standards. Whole genome sequencing was performed to genotype strains and classify the viral variants (see supplementary appendix subsection S7).

### Outcomes

The primary outcome measure was the rate of viral clearance estimated from viral genome densities in serial duplicate oropharyngeal viral swab eluates taken daily between days 0 and 7 (see statistical methods below and supplementary appendix S9 for the method of estimation). Secondary endpoints were:

i. all-cause admission to hospital for clinical deterioration (until day 28);
ii. time-to-resolution of fever in patients febrile at admission;
iii. time-to-resolution of symptoms.

These endpoints were assessed using survival methods because the data at the last visit were right-censored. Patients were defined as febrile at admission if at least one axillary temperature measurement within 24 hours of randomisation was ≥37.5°C. Resolution of fever was defined as an axillary temperature ≤37.0°C for at least 24 hours. Symptom resolution was defined as no reported symptoms. All adverse events were graded as per the Common Terminology Criteria for Adverse Events version 5.0.^28^ Summaries were generated if the adverse event was grade 3 or worse, and was new or had increased in intensity. Serious adverse events were recorded separately and reported to the data safety monitoring board, however there were no serious adverse events during this portion of the trial.

### Sample size and analysis framework

For each intervention, the sample size was adaptive, based on the prespecified futility and success stopping rules. A maximum sample size of 120 patients was prespecified (this does not include the no study drug arm or the positive control arm - currently ritonavir-boosted nirmatrelvir). Sample size requirements and thresholds for stopping rules were determined by simulation (see statistical analysis plan supplementary appendix subsection S9).

The primary outcome measure, the rate of viral clearance between day 0 and day 7, was expressed as a slope coefficient and estimated under a Bayesian hierarchical linear model with random effect terms for the individual patient slope and intercept.^25,29^ The model was fitted to the daily log_10_ oropharyngeal swab eluate viral densities (genomes/mL) between days 0 and 7 (18 measurements per patient), using weakly informative priors and treating non-detectable viral loads (CT value ≥40) as left-censored (supplementary appendix subsection S9).^25^ The treatment effect was defined as the multiplicative change (%) in the viral clearance rate, either relative to the no study drug arm (when determining if an intervention had an antiviral effect), or relative to the positive control arm (ritonavir-boosted nirmatrelvir).^29^ The viral clearance rate (i.e., slope coefficient from the model fit) can also be expressed as a clearance half-life (t_1/2_ = log_10_ 0.5/slope). A 50% increase in clearance rate equals a 33% reduction in clearance half-life. All models include as covariate terms on the slope coefficient the time since study commencement, the virus variant, and the study site.

Because of the changing pattern of viral variants, and the substantial increase in the rate of viral clearance since the beginning of the pandemic, each of the studied interventions was compared only against the concurrent controls, with interim analyses planned every additional ten patients recruited into each group. However, in practice, the interim analyses were less frequent than planned as recruitment occurred quickly. At first, all interim analyses compared the new intervention against the no study drug group. The protocol stipulated dropping the intervention for futility when there is >90% probability that it accelerated viral clearance by less than 20% (this threshold was increased from 12.5% in January 2023; statistical analysis plan version 3.0). If the new intervention reaches the success threshold (i.e., >90% probability it accelerated viral clearance >20% relative to no study drug), it is then compared with the positive control. This secondary comparison terminates when the intervention is shown to be inferior, non-inferior, or superior to the positive control group using a 10% non-inferiority margin. If the intervention is superior, it then replaces the positive control group. All stopping decisions are made using data from contemporaneously randomly assigned patients only.

All efficacy analyses were done in a modified intention-to-treat (mITT) population, comprising all patients with >2 days follow-up data. Safety data were analysed in all patients who had received ≥ one dose of the study drug. A sensitivity analysis was performed using a non-linear model fitted to the serial viral densities, which allows for an initial increase followed by a log-linear decrease (Supplementary appendix subsection S9).

### Additional post hoc analyses

A recent analysis of all available PLATCOV trial unblinded data (N>1200 patients) characterised the substantial increase in viral clearance rates that has occurred since the beginning of the platform trial 28 months ago. The average oropharyngeal viral clearance half-life in the no study drug arm has shortened from ~17 hours in late 2021 to ~9 hours in October 2023.^30^ This analysis also showed that restricting the primary endpoint to the clearance rate estimated over the first 5 days, instead of 7 days, resulted in greater power to assess treatment effects (i.e. larger z-scores between effective and ineffective or no drug arms). A post-hoc analysis of the fluoxetine data was therefore added in which the estimation of the viral clearance rates was made from the first 5 days only.

### Meta-analysis

To calibrate the effect sizes observed for the fluoxetine arm, an individual patient data meta-analysis was conducted of all small molecule drugs and monoclonal antibodies with unblinded data from the PLATCOV trial (favipiravir, remdesivir, molnupiravir, ritonavir-boosted nirmatrelvir, casirivimab/imdevimab and ivermectin). Not all interventions were randomised concurrently, so the time since study commencement was included as a covariate on the mean slope parameter to control for temporal confounding.

### Statistical analysis

All data analysis was done in R version 4.3.2. Posterior distributions were approximated using Hamiltonian Monte Carlo in Stan via the RStan interface.^31^ 4,000 iterations were run over four independent chains with 2000 iterations for burn-in. Convergence was assessed visually from the trace plots (Figures S4 and S5) and using the R-hat statistic (a value <1.1 was considered acceptable convergence). Goodness of fit was assessed by plotting the residuals over time and comparing the daily median model predictions with the observed values (Figure S5). All point estimates are reported with 95% credible intervals (CrIs), defined by the 2.5% and the 97.5% quantiles of the posterior distribution. Model fits were compared using approximate leave-one-out comparison as implemented in the package loo.^32^

## Results

The PLATCOV platform trial began recruitment on 30^th^ September 2021. The fluoxetine arm was added in Thailand on 5^th^ April 2022, and in the other sites on the 21^st^ June 2022, and was stopped on 8^th^ May 2023 after 120 patients had been randomised to fluoxetine. Of the 675 patients randomised during that period, 120 patients were randomised to fluoxetine, 151 to no study drug, and the remaining 404 were randomised to other interventions (casirivimab/imdevimab, tixagevimab/cilgavimab, nitazoxanide, favipiravir, remdesivir, ivermectin, ensitrelvir, ritonavir-boosted nirmatrelvir and molnupiravir) (Figure 1). Four patients from the fluoxetine group withdrew consent. Two patients from the no study drug arm withdrew consent.

**Figure 1:**
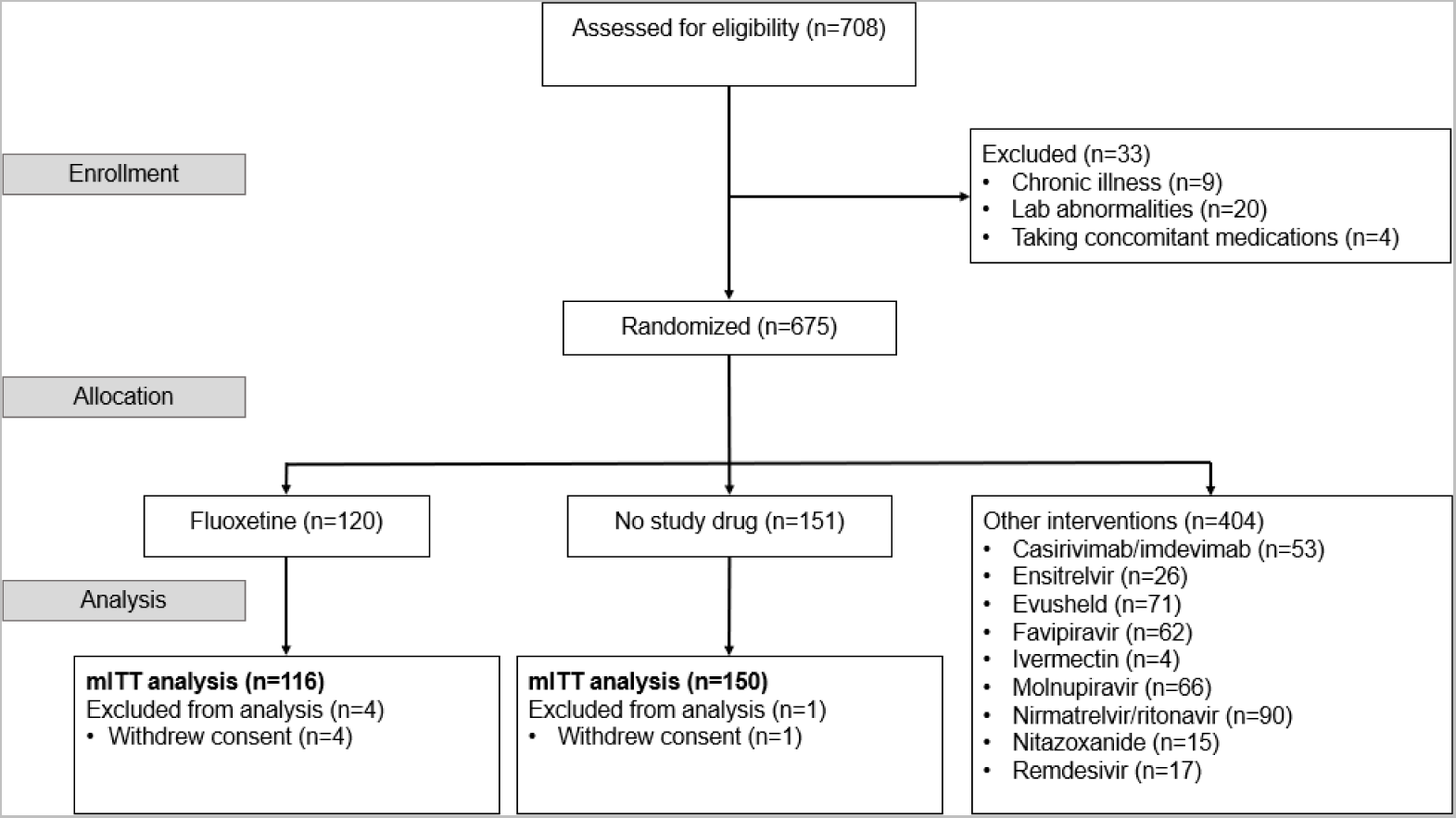
Study CONSORT diagram for the fluoxetine versus no study drug analysis. *In Thailand, pre-screening occurred in the Acute Respiratory Infection (ARI) unit of the Hospital for Tropical Diseases, Bangkok. Potentially eligible patients were selected by the ARI Nurses to be contacted by the study team. Therefore, a high proportion of those assessed for eligibility participated in the study.

The majority of patients (89.6%) were enrolled in Bangkok, Thailand (Table 1). The median interval since symptom onset was 2 (IQR: 2 to 3) days. Most patients had high oropharyngeal eluate viral densities at presentation; average viral density of ~350,000 genomes per mL. Patients were infected with a wide variety of virus variants, the 3 most common being BA.2.75 (107/355), BA.5 (99/355) and XBB.1.5-like (63/355).

**Table 1:** Admission patient characteristics in the mITT population. For categorical variables, the number (%) is shown and for continuous variables the standard deviation.

|  | No study drug | Fluoxetine |
| --- | --- | --- |
| All sites | 150 | 116 |
| Brazil | 17 (11.3%) | 12 (10.3%) |
| Thailand | 129 (86.0%) | 101 (87.1%) |
| Laos | 0 (0%) | 1 (0.9%) |
| Pakistan | 4 (2.7%) | 2 (1.7%) |
| Age (years) (SD) | 30.5 (7.8) | 29.5 (7.7) |
| Female N (%) | 98 (65.3%) | 82 (70.7) |
| Weight (kg) (SD) | 62.7 (13.4) | 59.6 (11.3) |
| Body mass index (kg/m<sup>2</sup>) (SD) | 23.2 (4.0) | 22.3 (3.5) |
| Baseline oropharyngeal eluate viral density<br>(log<sub>10</sub> copies per mL) (SD) | 5.5 (1.4) | 5.6 (1.3) |
| Symptom onset (days) (SD) | 2.1 (0.8) | 2.2 (0.8) |
| Vaccinated (%) | 150 (100.0%) | 116 (100.0%) |
| SARS-CoV-2 variants |  |  |
| BA.2 (%) | 30 (20.0%) | 24 (20.7%) |
| BA.2.3.20 (%) | 0 (0.0%) | 1 (0.9%) |
| BA.2.75 (%) | 41 (27.3) | 34 (29.3%) |
| BA.4 (%) | 2 (1.3%) | 0 (0.0%) |
| BA.5 (%) | 42 (28.0%) | 31 (26.7%) |
| BN.1.9 (%) | 2 (1.3) | 0 (0.0%) |
| XBB (%) | 10 (6.7%) | 9 (7.8%) |
| XBB.1.5-like (%) | 23 (15.3%) | 15 (12.9%) |
| Others (%) | 0 (0.0) | 2 (1.7) |

### Tolerability

The oropharyngeal swabbing procedures and all treatments were well-tolerated. Patients allocated to the fluoxetine arm reported increased somnolence compared to the no study drug arm, and so the treatment was given in the evening.

### Clinical responses

There were no serious adverse events (SAEs), hospitalisations or deaths in either arms, and no patients developed severe disease. There were no significant differences in times to symptom resolution or fever clearance between the fluoxetine and the no study drug arms, however this latter comparison had low power as only a quarter of patients were febrile at baseline (supplementary appendix figures S6 and S7).

### Virological responses

Rates of viral clearance were estimated in the mITT population (6,362 measurements in 355 patients, of which 5,062 (80%) were above the lower limit of quantification). Under the linear model fluoxetine resulted in a 15% (95% CrI: 2 to 34%) faster average rate of viral clearance over 7 days relative to no study drug. (Figure 2). The posterior probability that the effect of fluoxetine was less than the pre-specified futility margin of 20% was 0.70. The non-linear model gave very similar estimates: an acceleration in viral clearance rate of 11% (95% CrI −3 to 29%) relative to the no study drug. Under the linear model, the median estimated viral clearance half-lives were 14.0 hours (9.3–18.0) with fluoxetine, and 14.9 hours (11.5–20.8) in the concurrent no study drug group (Figure 3).

**Figure 2:**
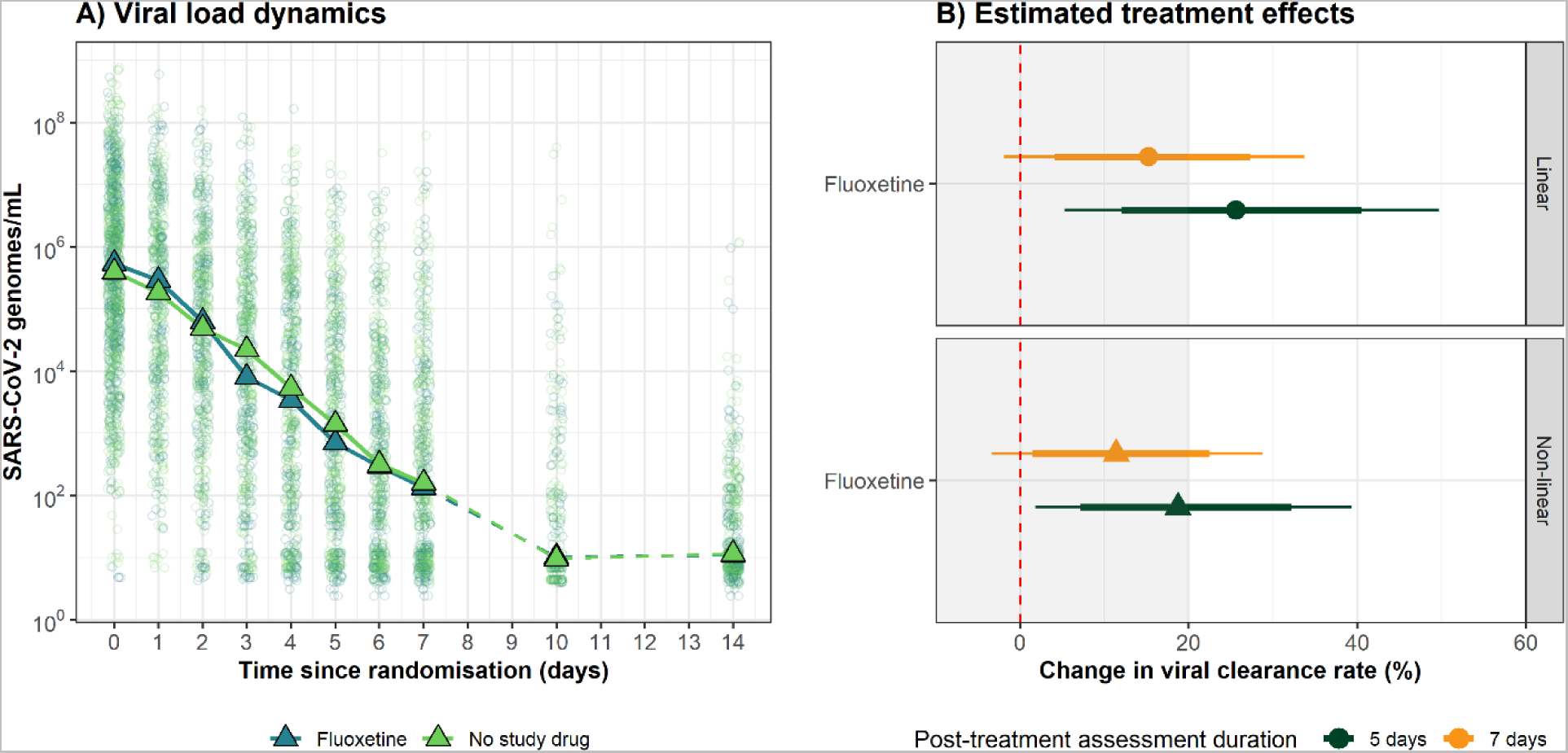
Antiviral effect of fluoxetine in early COVID-19. Panel A: individual viral densities data (fluoxetine: dark green; no study drug: light green). Triangles show the daily median oropharyngeal eluate viral densities by arm. Panel B: posterior estimates of the treatment effects of fluoxetine relative to no study drug, under the linear and non-linear models (orange: viral clearance assessed over 7 days; green: viral clearance assessed over 5 days). Thick and thin error bars represent the 80% and 95% CrIs, respectively. The shaded area indicates the prespecified futility zone (<20% increase in viral clearance rate).

**Figure 3:**
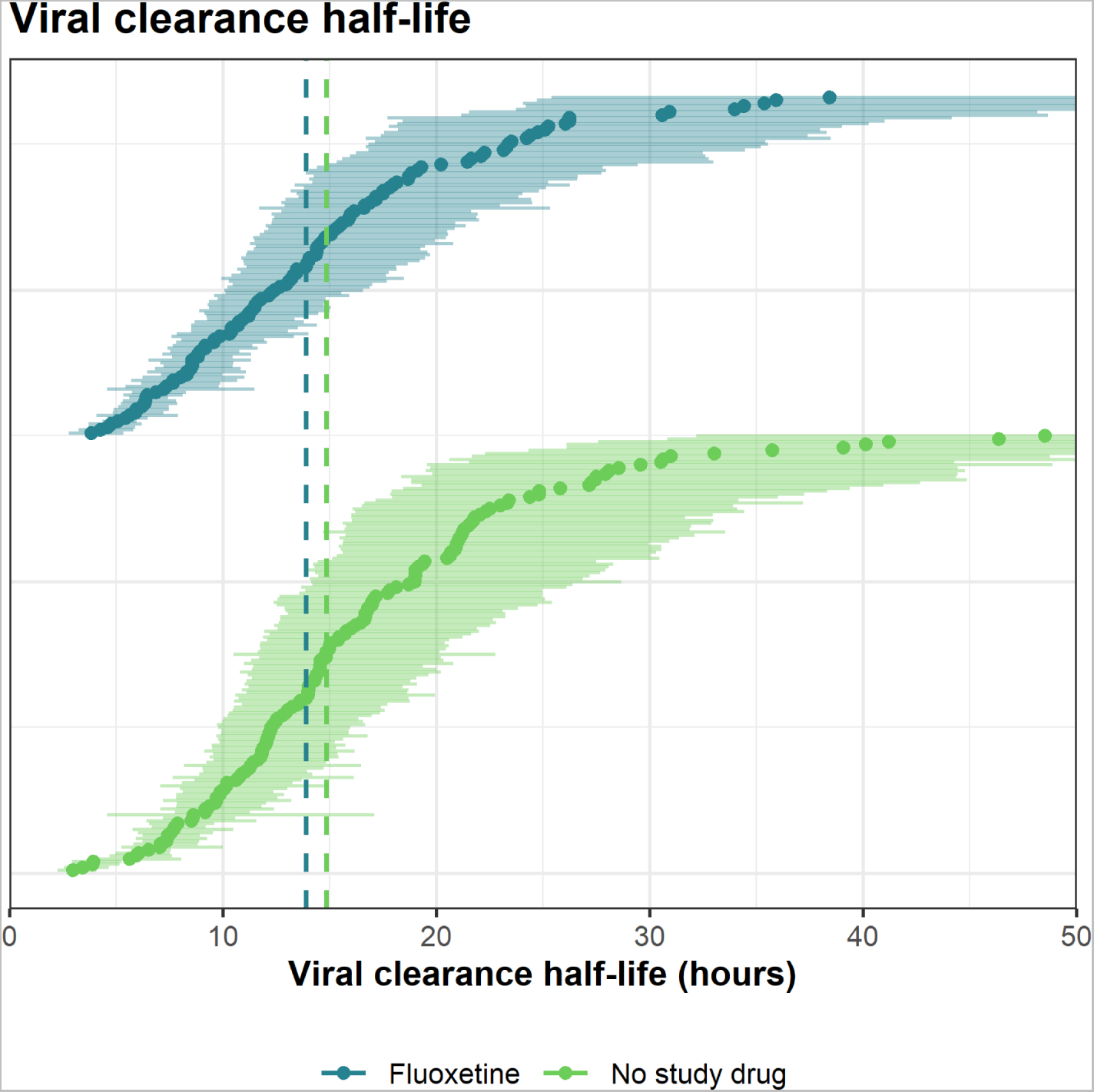
Estimated SARS-CoV-2 clearance half-lives (in hours) estimated over 7 days for individual patients in the fluoxetine arm (dark green), and the no-study-drug arm (light green). The median estimates (circles) and 80% credible intervals (error bars) are displayed. Vertical dashed lines indicate the median half-lives of each group.

An in-depth analysis of all unblinded data (not including the fluoxetine arm) from the platform trial showed that viral clearance rates have increased substantially since the study began in September 2021.^30^ This analysis indicated that maximal power to detect an antiviral effect was obtained from clearance rates estimated over the first 5 days (rather than 7 days). As a *post hoc* sensitivity analysis, we therefore estimated the treatment effect of fluoxetine using data only from the first 5 days after randomisation. Under the linear and non-linear models, the estimated fluoxetine treatment effects were substantially larger: 26% (95% CrI: 5 to 50%) under the linear model; and 18% (95% CrI: 2 to 39%) under the non-linear model (Figure 2B).

### Meta-analysis

Under the linear model analysing viral clearance rates over 7 days, the meta-analysis including all unblinded drugs (not concurrently randomised) and adjusting for calendar time and viral variants, estimated that fluoxetine increased viral clearance by 16% (95% CrI 3 to 32%) compared to the no study drug group (Figure 4). Fluoxetine treatment resulted in a higher viral clearance rate than two interventions previously reported to have no clinical antiviral effect; ivermectin^25^and favipiravir.^27^ The treatment effect of fluoxetine was lower than that of casirivimab/imdevimab,^26^ remdesivir,^26^ molnupiravir,^2^ and substantially lower than ritonavir-boosted nirmatrelvir,^2^ with the probabilities of 0.88, 0.94, 0.98, and 1.00, respectively. These four active antivirals/monoclonal antibodies increased the rates of viral clearance by 29% (95% CrI 10 to 48%), 35% (95% CrI 14 to 59%), 37% (95% CrI 18 to 60%), and 85% (95% CrI 61 to 112%), respectively. Additionally, consistent with the main analysis, a post-hoc analysis of viral clearance estimated over 5 days demonstrated higher discriminating power (figure 4B) indicated that fluoxetine increased the viral clearance rate by 28% (95% CrI 11 to 49%).

**Figure 4:**
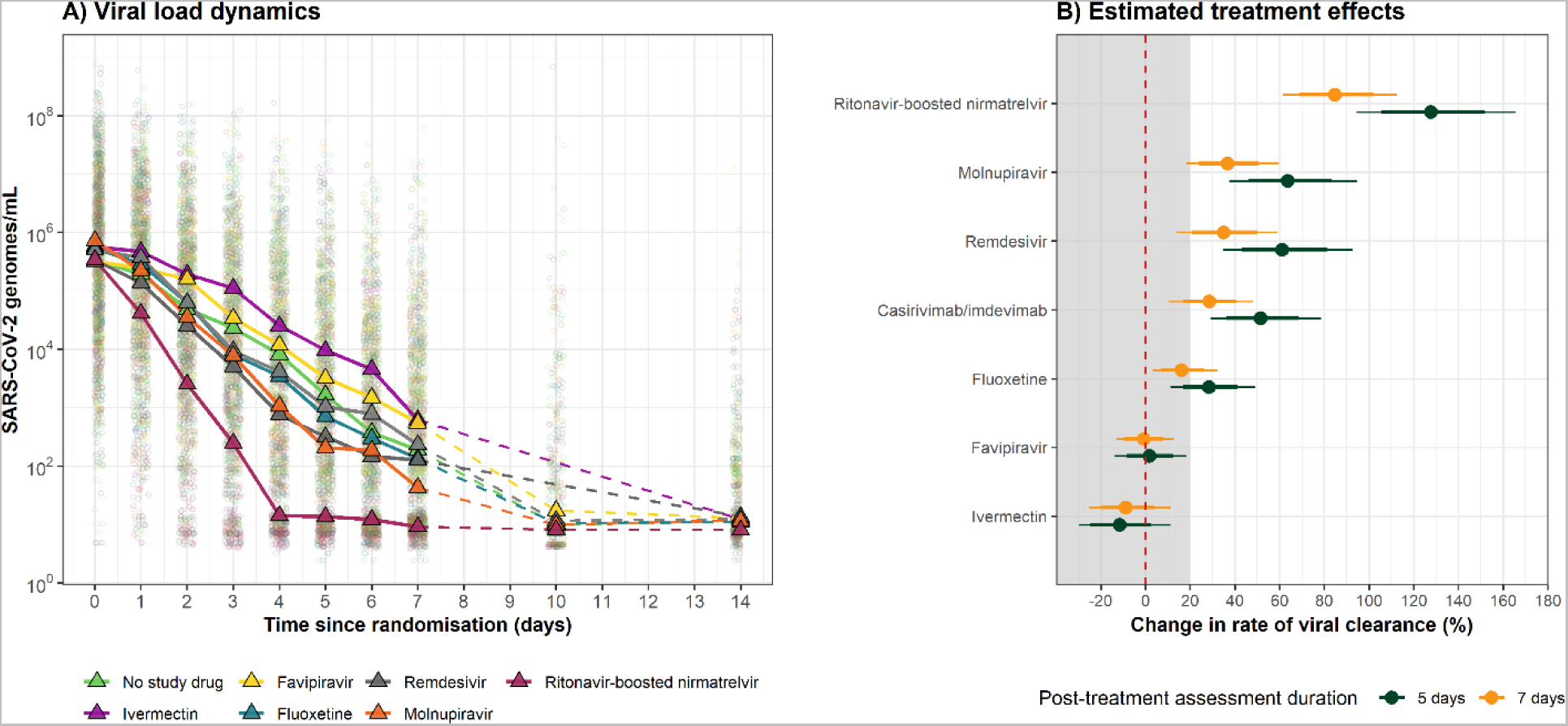
Meta-analysis of oropharyngeal viral clearance rates in 783 patients in unblinded drug arms (ritonavir-boosted nirmatrelvir, molnupiravir, remdesivir, casirivimab/imdevimab, fluoxetine, favipiravir, and ivermectin relative to no study drug; not all concurrently) (A) Daily median oropharyngeal viral loads by treatment group. (B) Posterior estimates of treatment effect on viral clearance rate relative to no study drug under a model adjusting for calendar time and virus variant (orange: viral clearance assessed over 7 days; green: viral clearance assessed over 5 days). Thick and thin error bars representing the 80% and 95% CrIs, respectively. The shaded area indicates the futility zone (<20% increase in viral clearance rate).

## Discussion

This clinical pharmacodynamic evaluation shows that the widely used SSRI fluoxetine, which has the same proposed FIASMA mechanism of action as fluvoxamine, does have measurable, albeit weak antiviral activity against SARS-CoV-2 *in vivo*. Earlier in the pandemic, before effective antivirals and before vaccines were deployed, any available drug with anti-SARS-CoV-2 activity could have played a role in the management of COVID-19. Based on several observational studies which reported lower mortalities in patients receiving SSRIs, ^4–7^ and some evidence for prophylactic activity,^8^ the SSRIs fluoxetine and fluvoxamine were proposed as treatments for early COVID-19. Both drugs are interesting choices as they are inexpensive, widely available, very widely used, and have excellent safety profiles. A meta-analysis of randomised controlled trials of fluvoxamine showed a slight, but non-significant reduction in hospitalisation +/- mortality (Supplementary appendix; subsection S1). However, these results were not sufficient to change treatment policies and practices. In May 2022, the US FDA rejected an emergency use authorisation (EUA) for fluvoxamine maleate in outpatients with COVID-19, on the basis that there was insufficient evidence that fluvoxamine can prevent progression to severe disease or hospitalisation. The US FDA noted that “it is unlikely that fluvoxamine possesses a high degree of activity against SARS-CoV-2”.^33^ There have been no randomised controlled trials in outpatients assessing fluoxetine.

In this comparative *in vivo* pharmacodynamic platform trial, carried out in low-risk adults with early symptomatic COVID-19 infection, fluoxetine demonstrated significant but weak antiviral activity. This was not sufficient for it to reach the prespecified success threshold of a 20% acceleration of viral clearance (assessed over 7 days) compared to the contemporaneous control group. This threshold was set because there are now highly effective antiviral drugs for early symptomatic COVID-19, and so it is unlikely that drugs with a substantially lower potency would be used in treatment. The main protease inhibitor nirmatrelvir, in combination with ritonavir, is currently the most effective antiviral treatment assessed in this platform trial. In the 7-day assessment its acceleration of viral clearance was over five times greater than that of fluoxetine. But it has several disadvantages. Ritonavir-boosted nirmatrelvir is very expensive and often unaffordable, it is not readily available worldwide, it produces an unpleasant taste, and ritonavir is contraindicated in many patient groups due to drug interactions. So, although fluoxetine is unlikely to be used in treatment now, it might still have a role in prevention where less antiviral activity is required than in the treatment of an already established infection (hence why lower doses are effective in prophylaxis, than in treatment).

The methodology used in this platform trial is an effective way to measure antiviral effects in COVID-19. Acceleration of viral clearance reflects the *in vivo* antiviral effect and correlates with prevention of hospitalisation and death.^34^ It is becoming increasingly difficult to carry out large trials with clinical endpoints. This is because the low rates of hospitalisation and death in COVID-19 infections with current viral variants in an increasingly immune population mean that sample sizes using these endpoints must be prohibitively large.^22^ Virological pharmacodynamic endpoints can be used to measure antiviral effects with substantially smaller sample sizes. The pharmacodynamic assessment has also become simpler. Increased rates of viral clearance since the pandemic started now mean that viral clearance can be measured more accurately over 5 rather than 7 days.

The study has several limitations. It is open-label, which may have influenced the symptom reporting in each arm. There is substantial variability in estimated serial viral densities and much of the inter-subject variance in viral clearance rates is unexplained. There still remains some uncertainty about the antiviral potency of fluoxetine (at the doses evaluated). Whether larger doses would have greater activity is not known, although tolerability would have been reduced. The applicability of this result to other SSRIs or other FIASMAs was not determined.

In summary, fluoxetine demonstrates modest *in vivo* antiviral activity in early COVID-19. It accelerated viral clearance by at least 15%, but this is much less than currently available effective antivirals. Given that there are more effective, albeit much more expensive drugs, fluoxetine is unlikely to be used in the treatment of COVID-19 at this stage of the pandemic, but whether it could have had a useful role earlier is unclear. Fluoxetine might have a role either in prophylaxis, or in high-risk patients unable to access or take other treatments, or in future pandemics, but further evidence will be needed before such recommendations can be made. *In vivo* pharmacodynamic assessments of drugs should be more widely adopted.

## Acknowledgements

The trial was generously supported by the Wellcome Trust Grant ref: 223195/Z/21/Z through the COVID-19 Therapeutics Accelerator.

We thank all the patients with COVID-19 who volunteered to be part of the study. We thank the data safety and monitoring board (DSMB) (Tim Peto, André Siqueira, and Panisadee Avirutnan); the trial steering committee (TSC) (Nathalie Strub-Wourgaft, Martin Llewelyn, Deborah Waller, and Attavit Asavisanu); Sompob Saralamba and Tanaphum Wichaita for developing the RShiny randomisation app; and Mavuto Mukaka for invaluable statistical support. We also thank all the staff of the Clinical Trials Unit (CTU) at MORU, PCR Expert group (Janjira Thaipadungpanit, Audrey Dubot-Pérès and Clare Ling), Thermo Fisher for their excellent support with this project, and all the hospital staff at the Hospital for Tropical Diseases, Faculty of Tropical Medicine, as well as those involved in sample processing in MORU and the processing and analysis at the Faculty of Tropical Medicine (FTM), molecular genetics laboratory. We would thank the MORU Clinical Trials Support Group (CTSG) for data management, monitoring, ethics and regulatory submissions and logistics, and the purchasing, administration, and support staff at MORU, and those at the Brazil site who provided expert help in managing patients (Joseane Fratari, Josiane Vaz and Fátima Brant). We would finally like to thank everyone who supported the Laos and Pakistan sites.

## Contributors

PJ-investigation, methodology, project administration, supervision, validation, and writing-original draft. SB-investigation, methodology, project administration, writing-original draft. PJ and SB contributed equally. WHKS-funding acquisition, investigation, methodology, project administration, supervision, validation, and writing-original draft. JAW-conceptualisation, data curation, formal analysis, funding acquisition, methodology, visualisation, and writing-original draft. TN, TS, VL-Investigation, methodology, supervision. EMB-data curation, formal analysis, visualisation. PW-data curation, formal analysis, visualisation, and writing-original draft. RA, FA, NG-formal analysis, investigation. LE, PA, CC, JJC, SS, VK, TN, JT, FQ, AMK - methodology, investigation, project administration. WM, KS, AP-investigation, methodology. BH, KP-methodology, investigation, supervision. MP, AS, BL-resources. WRJT-methodology, supervision. KC, MI-formal analysis, investigation, resources, supervision. SP, AMD, AB, MMT, WP, WP, DC, SV - methodology, investigation, resources, supervision. NPJD-funding acquisition, methodology, investigation, resources, supervision. NJW-conceptualisation, funding acquisition, methodology, supervision, validation, and writing-original draft.

All Authors were involved in writing-review & editing.

JAW, WHKS, EMB, TN, MI and NJW have directly accessed and verified the underlying data reported in the manuscript.

## Data sharing statement

All code and de-identified participant data required for replication of the study’s endpoints are openly accessible via GitHub, as well as the study protocol and statistical analysis plan, from publication date onwards: https://github.com/jwatowatson/PLATCOV-Fluoxetine Individual Patient Data can be requested and may be shared according to the terms defined in the MORU data sharing policy with other researchers to use in the future from the date of publication. Further information on how to apply is found here: https://www.tropmedres.ac/units/moru-bangkok/bioethics-engagement/data-sharing.

