## Supplementary Materials for "Antiviral efficacy of fluoxetine in early symptomatic COVID-19: an open-label, randomised, controlled, adaptive platform trial (PLATCOV)"

### S1: List of Sites and Investigators (PLATCOV Collaborative Group)

#### Sites

1. Hospital for Tropical Diseases (HTD), Faculty of Tropical Medicine, Mahidol University, 420/6 Rajvithi Road, Bangkok, 10400, Thailand
2. Universidade Federal de Minas Gerais, Av. Antônio Carlos, 6627 Belo Horizonte, Minas Gerais 31270 – 901, Brazil
3. Mahosot Hospital, Quai Fa Ngum, Vientiane, Laos
4. Aga Khan University, National Stadium Rd, Karachi, Pakistan

**Co-principal investigators:**

Nicholas J White[1,2]

William HK Schilling[1,2]

**Faculty of Tropical Medicine, Mahidol University:**

**Site and Country Principal investigator:**

Weerapong Phumratanaprapin[3]

**Accountable Investigator:**

Viravarn Luvira[3]

**Co-Investigators/team members:**

James J Callery[1,2]

Nicholas PJ Day[1,2]

Sasithon Pukrittayakamee[1,3]

Simon Boyd[1,2]

Cintia Cruz[1,2]

Arjen M Dondorp[1,2]

Walter RJ Taylor[1,2]

James A Watson[1,4]

Phrutsamon Wongnak [1,2]

Watcharapong Piyaphanee[3]

Kittiyod Poovorawan[1,3]

Thundon Ngamprasertchai[3]

Tanaya Siripoon[3]

Borimas Hanboonkunupakarn[1,3]

Kesinee Chotivanich[1,3]

Podjanee Jittamala[1,5]

Mallika Imwong[1,6]

Maneerat Ekkapongpisit[1]

Varaporn Kruabkontho[1]

Thatsanun Ngernseng[1]

Jaruwan Tubprasert[1]

Mohammad Yazid Abdad[1,2]

Srisuda Keayarsa[3]

Orawan Anunsittichai[1]

Maliwan Hongsuwan[1]

Yutatirat Singhaboot[3]

Wanassanan Madmanee[1]

Elizabeth M Batty[1,2]

Runch Tuntipaiboontana[1]

Watcharee Pagornrat[1]

Amornrat Promsongsil[1]

Shivani Singh[1,2]

Manisaree Saroj[1]

Jindarat Kouhathong [1]

Kanokon Suwannasin[1]

Ellen Beer[1]

Tanatchakorn Asawasriworanan[1]

Stuart Blacksell[1,2]

Salwaluk Panapipat[1]

Naomi Waithira[1,2]

Joel Tarning[1,2]

Nuttakan Tanglakmankhong[1]

**Bangplee Hospital: (**discontinued before Fluoxetine assessment)

**Site Principal investigator:**

Pongtorn Hanboonkunupakarn [6]

**Co-investigator:**

Sakol Sookprome[6]

**Vajira Hospital:** (discontinued before Fluoxetine assessment)

**Site Principal investigator:**

Vasin Chotivanich[8]

**Co-investigators:**

Wiroj Ruksakul[8]

Chunlanee Sangketchon[9]

**Brazil Site – Universidade Federal de Minas Gerais**

**Site and Country Principal investigator:**

Mauro M Teixeira[10]

**Co-Investigators:**

Lisia M Esper[10]

Fernando R Ascencao[11]

Renato S Aguiar[12]

Pedro J Almeida[10]

**Laos Site – Mahosot Hospital:**

**Site Principal investigator:**

Elizabeth Ashley [2,13]

**Co-Investigators:**

Audrey Dubot-Pérès[2,13, 14]

Mayfong Mayxay [2,13,15]

Manivanh Vongsouvath[16]

Danoy Chommanam[13]

Latsaniphone Boutthasavong [13]

Vayouly Vidhamaly[13]

Koukeo Phommasone[13]

Terry John Evans[2,13]

Susath Vongphachanh [17]

Sisouphanh Vidhamaly[18]

Ammala Chingsanoon [18]

Sixiong Bisayher [18]

**Pakistan Site – Aga Khan Hospital:**

**Site and Country Principal investigator:**

M Asim Beg [19]

**Co-Investigators:**

Abdul Momin Kazi[19]

Farah Qamar[19]

Najia K Ghanchi[19]

Syed Faisal Mahmood [19]

**Thailand Ministry of Public Health:**

Manus Potaporn[20]

Attasit Srisubat[20]

Bootsakorn Loharjun [20]

**Affiliations:**

1. Mahidol Oxford Tropical Medicine Research Unit, Faculty of Tropical Medicine, Mahidol University, Bangkok, Thailand

2. Centre for Tropical Medicine and Global Health, Nuffield Department of Medicine, Oxford University, Oxford, UK

3. Department of Clinical Tropical Medicine, Faculty of Tropical Medicine, Mahidol University, Bangkok, Thailand

4. Oxford University Clinical Research Unit, Vietnam

5. Department of Clinical Tropical Hygiene, Faculty of Tropical Medicine, Mahidol University, Bangkok, Thailand

6. Department of Molecular Tropical Medicine and Genetics, Faculty of Tropical Medicine, Mahidol University, Bangkok, Thailand

7. Bangplee Hospital, Ministry of Public Health, Samut Prakarn province, Thailand

8. Faculty of Medicine, Navamindradhiraj University, Bangkok, Thailand

9. Faculty of Science and Health Technology, Navamindradhiraj University, Bangkok, Thailand

10. Clinical Research Unit, Center for Advanced and Innovative Therapies, Universidade Federal de Minas Gerais, Brazil

11. Department of Biochemistry and Immunology, Universidade Federal de Minas Gerais, Brazil

12. Department of Genetics, Ecology and Evolution, Institute of Biological Sciences, Universidade Federal de Minas Gerais, Brazil

13. Lao-Oxford-Mahosot Hospital-Wellcome Trust Research Unit, Microbiology Laboratory, Mahosot Hospital, Vientiane, Lao P.D.R.

14. Unité des Virus Émergents, Marseille, France

15. Institute for Research and Education Development, University of Health Sciences, Vientiane, Lao P.D.R.

16. Microbiology Laboratory, Mahosot Hospital, Vientiane, Lao P.D.R.

17. Mahosot Hospital, Vientiane, Lao P.D.R.

18. Pulmonology Department, Mahosot Hospital, Vientiane, Lao P.D.R.

19. Aga Khan University, Karachi, Pakistan

20. Department of Medical Services, Ministry of Public Health, Nonthaburi, Thailand

#### S2: Ethics Approval

The trial was approved by local and national research ethics boards in Thailand (Faculty of Tropical Medicine Ethics Committee, Mahidol University, FTMEC Ref: TMEC 21-058) and the Central Research Ethics Committee (CREC, Bangkok, Thailand, CREC Ref: CREC048/64BP-MED34), in Brazil by the Research Ethics Committee of the Universidade Federal de Minas Gerais (COEP-UFMG, Minas Gerais, Brazil, COEP-UFMG) and National Research Ethics Commission- (CONEP, Brazil, COEP-UFMG and CONEP Ref: CAAE:51593421.1.0000.5149), in Laos by the National Ethics Committee for Health Research (NECHR, Lao People's Democratic Republic, Submission ID 2022.48) and the Federal Drug Administration (FDA, Lao People's Democratic Republic, 13066/FDD_12Dec2022), in Pakistan by the National Bioethics Committee (NBC No.4-87/COVID-111/22/842) the Ethics Review Committee (ERC 2022-7496-21924) and the Drug Regulatory Authority (DRAP Ref: No.03-18/2022-CT (PS)) and finally by the Oxford University Tropical Research Ethics Committee (OxTREC, Oxford, UK, OxTREC Ref: 24-21).

#### S3: Baseline procedures

Baseline investigations included a full clinical examination, rapid SARS-CoV-2 antibody test (BIOSYNEX COVID-19 BSS IgM/IgG®, Illkirch-Graffenstaden, France, done in Thailand only), blood sampling for haematology and biochemistry, an electrocardiogram, and a chest radiograph (following local guidance in Thailand, but not a study requirement).

#### S4: Symptoms included in symptom questionnaire performed during study visits

- Fever
- Headache
- Dizziness
- Blurred vision
- Fatigue
- Cough
- Difficulty breathing
- Chest pain
- Runny nose
- Loss of smell or taste
- Abdominal pain
- Loss of appetite
- Nausea
- Vomiting
- Diarrhoea
- Arthralgia
- Myalgia
- Itching
- Skin rash
- Sore throat
- Other

S5: Adverse events (AE) for fluoxetine

Supplementary table 1: Summary of adverse events (grade 3 and above) for fluoxetine

| | All grades | | Grade 3-4 | |
|---|---|---|---|---|
| | **Fluoxetine (n=0)** | **No study drug (n=0)** | **Fluoxetine (n=0)** | **No study drug (n=0)** |
| Any adverse event (Grade ≥ 3) | 0 | 0 | | |
| Serious adverse event reported | 0 | 0 | | |
| Symptoms | | | | |
| Fever | | | 0 | 0 |
| Headache | | | 0 | 0 |
| Dizziness | | | 0 | 0 |
| Blurred vision | | | 0 | 0 |
| Fatigue | | | 0 | 0 |
| Cough | | | 0 | 0 |
| Difficulty breathing | | | 0 | 0 |

| | | | | |
|---|---|---|---|---|
| Chest pain | | | 0 | 0 |
| Runny nose | | | 0 | 0 |
| Loss of smell or taste | | | 0 | 0 |
| Abdominal pain | | | 0 | 0 |
| Loss of appetite | | | 0 | 0 |
| Nausea | | | 0 | 0 |
| Vomiting | | | 0 | 0 |
| Diarrhoea | | | 0 | 0 |
| Arthralgia | | | 0 | 0 |
| Myalgia | | | 0 | 0 |
| Itching | | | 0 | 0 |
| Skin rash | | | 0 | 0 |
| Laboratory abnormalities | | | | |
| Creatinine | | | 0 | 0 |
| BUN | | | 0 | 0 |
| Sodium | | | 0 | 0 |
| eGFR | | | 0 | 0 |
| Potassium | | | 0 | 0 |

| | | | | |
|---|---|---|---|---|
| ALT/SGPT | | | 0 | 0 |
| AST/SGOT | | | 0 | 0 |
| Total bilirubin | | | 0 | 0 |
| Direct bilirubin | | | 0 | 0 |
| Alkaline Phosphatase | | | 0 | 0 |
| LDH | | | 0 | 0 |
| Creatinine phosphokinase (CPK) | | | 0 | 0 |
| Anemia | | | 0 | 0 |
| Leukocytopenia | | | 0 | 0 |
| Neutropenia | | | 0 | 0 |
| Thrombocytopenia | | | 0 | 0 |

*All AEs solicited were reported between days 0-7, day 10, day 14 and at day 28.*

#### S6: Serious Adverse Events

There were no contemporaneous serious adverse events (SAEs) in the no study drug arm and none in the fluoxetine arm.

### S7: Virus variant determination

#### Brazil site – Virus variant determination

***SARS-CoV-2 whole-genome sequencing***

The virus sequencing was carried out using two different technologies, Illumina (Illumina, USA) and IonTorrent (ThermoFisher Scientific, USA). Only SARS-CoV-2-positive samples with Ct < 30 values for virus targets were considered. Illumina libraries were prepared using the QIAseq FX DNA Library Prep kit® (QIAGEN, Germany) and sequenced on the Illumina MiSeq® platform (Illumina, USA) with a v3 (600 cycles) cartridge, following all the manufacturer's protocols. IonTorrent libraries were prepared using the Ion AmpliSeq SARS-CoV-2 Panel® (ThermoFisher Scientific, USA) and sequenced on the IonTorrent PGM platform® with a 314-chip kit (ThermoFisher Scientific, USA), according to the manufacturer's recommendations. Three negative controls were used in all sample processing steps (cDNA synthesis, viral genome amplification, and library preparation).

***Viral genome assembly and classifications***

A custom pipeline was used to process the sequencing data. In the first step, quality control was performed with Trimmomatic v0.39. Adapter and primer sequences, short reads (< 50 nucleotides), and low-quality bases (Phred score < 30) were removed. Next, reads were mapped against the SARS-CoV-2 reference genome (GenBank accession: NC_045512) with Bowtie2. Samtools manipulated the mapping files, whilst consensus genome sequences were estimated using the bcftools consensus option. Masking of low-coverage sites was performed with bedtools. The code for the described pipeline can be found on GitHub (https://github.com/filiperomero2/ViralUnity). Depth thresholds differed between sequencing technologies employed. For IonTorrent data, sites with less than 20-fold depth were masked, while for Illumina, the minimum threshold was 10-fold. Sequences <70% genome coverage breadth were removed from downstream analysis.

#### Thailand and Laos sites – Virus variant determination

##### ***SARS-CoV-2 whole-genome sequencing***

The sequencing method carried out in this experiment follows the "PCR tiling of SARS-CoV-2 virus with rapid barcoding and Midnight RT PCR Expansion" provided by Oxford Nanopore Technology (Oxford, UK) developed based on a protocol by the ARTIC network group[1] and Freed *et al.*[2] Library preparation process started with reverse transcription, which consists of mixing the purified viral RNA with LunaScript RT SuperMix and incubating the mixtures in a thermal cycler. DNA fragments used in the assembly process were amplified by PCR using Midnight primer set (V3) and attached with barcodes from Rapid Barcode Plate (RB96). The mixtures from each sample were pooled together, cleaned with AMPure XP Beads (AXP), and attached with Rapid Adapter F (RAP F). The prepared DNA fragments were then loaded into a primed flow cell (FLO-MIN106) and sequenced on GridION MK1 system (MinION Mk1B system for Laos).

##### ***Viral genome assembly and classification***

The output sequencing data (.fast5) from MinKNOW software was base-called with Guppy software using the High Accuracy (HAC) model to generate nucleotide sequence data for each fragment (reads) in the fastq format. These base-called data were then processed through the established workflow wf-artic on EPI2ME software to be assembled into consensus sequences. Only reads with average Phred Quality (Q) score above 9 and minimum and maximum length of 250 and 1500 bps were used in the assembly process.

For viral classification - Consensus sequences from all sites were classified using the Pangolin tool (4.1.1) and Pangolin dataset (v1.14). Viral lineages were classified into eight categories corresponding to current and previous Variants of Concern (VOC): Delta, BA.1, BA.2, BA.2.75, BA.4, BA.5, XBB, and XBB.1.5-like. A lineage was classified as XBB.1.5-like based on the ECDC listing of Variants of Concern and includes lineages classified as XBB.1.5-like+F456L. All other XBB sublineages were classified as XBB.[3]

### S8: Randomization

The randomisation sheets were generated by the trial statistician (James Watson).

All new randomisation sheets and all updates of existing randomisation sheets were done using a pre-written R script which was stored on the randomisation Dropbox folder (owner is MORU, under custodianship of the head of MORU IT); this file is a full 'Professional' version with history recorded and only the trial statistician and head of IT had access to it. The file took the following inputs:

- Site codes (e.g. "th001") for which to generate randomisation sheets;
- The set of arms available for randomisation in that site;
- The number of arm repeats per block (this is set to the minimum integer such that in each block there is an integer number for each arm);
- The randomisation data file from each site (which has the patient numbers for subjects already randomised) named data-XXX.csv (where XXX is the site code), if this does not yet exist a blank .csv (headers only) is generated.

This R script is run every time a new site becomes active and every time the set of available arms changes. The output is a .csv file named rand-XXX.csv (where XXX is the site code). This overwrites the pre-existing file (which can be retrieved from the Dropbox version® history). Each time the randomisation script is run, this is recorded on a log file.

The randomisation is done according to the following constraints:

- Blocks of 2*number of available arms;

- Additional ‘fuzziness’ by swapping one patient allocation per block at random (this can be swapped for any of the available arms) – this avoids knowing which arm the last patient per block will receive.

Each time an authorised member of the study team logs onto the web-app this is logged (timestamp and username).

Each time a new patient is randomised this is logged on to the file data-XXX.csv (where XXX is the site code) with the following information:

- Subject number
- Screening number
- Age
- Sex
- Member of study team username
- Timestamp

#### S9: Statistical Analysis

The primary analysis consists of fitting Bayesian hierarchical (mixed effects) linear models to the serial $\log_{10}$ viral load data up until day 7 (the day 14 data were not used in this analysis). All models encode residual error as a *t*-distribution with degrees of freedom estimated from the data. The *t*-distribution was chosen for robustness as the residual error is clearly non-Gaussian.[4,5] The *t*-distribution error model also makes the inferences robust against model misspecification (particularly for the linear models).[6] All models include correlated individual random effect terms for both the intercept (baseline viral load) and the slope. All changes to the slope are defined as multiplicative changes on the log scale (i.e. a value of 0 equals no change).

The treatment effect is defined as the proportional change (expressed as a multiplicative term) in the population slope of the daily change in $\log_{10}$ viral load (i.e. the decline in $\log_{10}$ viral load versus time). The data are modelled on the $\log_{10}$ copies per mL scale, after conversion from Ct values using the standard curve generated from the 12 control concentrations (synthetic samples with known viral densities) from each 96 well plate. The standard curve transformation is done by fitting a linear mixed effects model (random slope and random intercept for each plate) to the control data: regressing the Ct values on the known log viral densities. This borrows information across plates and adjusts for batch effects.

For all models, we adjusted the intercept and slope for the enrolling site (6 sites in total for all drugs, the reference site is the Hospital of Tropical Diseases which recruited >80% of patients) and for the virus variant called (Delta is reference: BA.1, BA.2, BA.5 are the alternatives). A subset of models also adjusted the slopes and intercepts for:

- Age
- Number of COVID-19 vaccine doses
- Days since symptom onset

All models adjust for human RNase P (proxy for the number of human cells in the sample). This is an independent linear predictor for each viral load measurement. We fitted the models using two sets of prior distributions: weakly informative priors (WIP) and non-informative priors (NIP).

***Models fitted***

For each analysis we fitted 8 separate models:

1. Model 1 is linear with RNase P adjustment; adjustment for site & variant; WIP. This is the main model used to report treatment effects.
2. Model 2 is non-linear; RNase P adjustment; adjustment for site & variant; WIP
3. Model 3 is linear with RNase P adjustment; adjustment for site, variant, age, days since symptom onset; WIP
4. Model 4 is non-linear with RNase P adjustment; adjustment for site, variant, age, days since symptom onset; WIP

Models 5 to 8 are the same as models 1-4 but with non-informative priors (NIP).

Model 1 was used for all stopping decisions. All models have RNase P adjustment and are all combinations of linear & non-linear models, with or without full covariate adjustment; and with either weakly informative priors or non-informative priors. We compared model fits using the loo (approximate leave-one-out cross validation) package. The statistical analysis plan provides a detailed overview of the model structures.

***Analyses***

Adaptive platform trials can suffer from temporal confounding. The analyses for the fluoxetine arm used concurrent controls only (i.e. patients in the no study drug arm who could have been randomized to the active arm). This included all controls enrolled up until the 8th May 2023.

***Code and analysis plan***

All data, models and analytical output are on the linked GitHub repository: https://github.com/jwatowatson/PLATCOV-Fluoxetine The Statistical Analysis Plan used (version 3.1) is also provided in this repository.

#### S10: Stopping rules

The stopping rules were determined using a simulation approach, based on previously modelled serial viral load data,[7] such that approximately 50 patients are needed to demonstrate increases in the rate of viral clearance of ~50%, with control of both type 1 and type 2 errors at 10%. The prespecified decision criteria for stopping a treatment arm were either a model-based probability of <0.1 that the intervention did not accelerate viral clearance by >12.5% relative to no study drug (futility), or >0.9 that it did (success). The first interim analysis (n=50) was prespecified as unblinded in order to review the methodology and stopping rules. Following this, the stopping threshold was increased from 5% to 12.5% because the treatment effect in the positive control arm (casirivimab/imdevimab) in SARS-CoV-2 Delta variant infections was substantially larger than expected. Thereafter trial investigators were blinded. The stopping threshold was later increased to 20%. Enrollment stopped when the DSMB approved the recommendation based on the data analysis.

The 5-day analyses were conducted and reported in the same way as the 7 days analysis.

### S11: Meta-analysis of small molecule drugs and monoclonal antibodies

Baseline characteristics of meta-analysis of small molecule drugs

| | No study drug | Ivermectin | Casirivimab /Imdevimab | Remdesivir | Favipiravir | Fluoxetine | Molnupiravir | Nirmatrelvir |
|---|---|---|---|---|---|---|---|---|
| All sites | 198 | 44 | 88 | 67 | 114 | 116 | 66 | 90 |
| Brazil | 25 (12.6%) | 0 (0.0%) | 0 (0.0%) | 9 (13.4%) | 16 (14.0%) | 12 (10.3%) | 0 (0.0%) | 0 (0.0%) |
| Thailand - FTM | 164 (82.8%) | 40 (90.9%) | 84 (95.5%) | 54 (80.6%) | 96 (84.2%) | 101 (87.1%) | 65 (98.5%) | 89 (98.9%) |
| Thailand – Vajira | 3 (1.5%) | 2 (4.5%) | 1 (1.1%) | 2 (3.0%) | 2 (1.8%) | 0 (0%) | 0 (0%) | 0 (0%) |
| Thailand – Bang Plee | 2 (1.0%) | 2 (4.5%) | 3 (3.4%) | 2 (3.0%) | 0 (0%) | 0 (0%) | 0 (0%) | 0 (0%) |
| Laos | 0 (0%) | 0 (0%) | 0 (0%) | 0 (0%) | 0 (0%) | 1 (0.9%) | 1 (1.5%) | 1 (1.1%) |
| Pakistan | 4 (2.0%) | 0 (0%) | 0 (0%) | 0 (0%) | 0 (0%) | 2 (1.7%) | 0 (0%) | 0 (0%) |
| Age (years) (SD) | 30.1 (7.6) | 30.0 (7.0) | 27.9 (7.3) | 30.1 (8.2) | 302 (7.5) | 29.5 (7.7) | 31.3 (7.5) | 30.3 (7.6) |
| Female N(%) | 128 (64.6%) | 24 (54.5%) | 55 (62.5%) | 35 (52.2%) | 71 (62.3%) | 82 (70.7%) | 37 (56.1%) | 57 (63.3%) |
| Weight (kg) (SD) | 62.8 (13.5) | 61.6 (12.3) | 60.4 (12.3) | 63.9 (11.0) | 63.0 (13.6) | 59.6 (11.3) | 63.4 (14.7) | 62.8 (12.3) |
| Body mass index ($kg/m^2$) (SD) | 23.1 (4.1) | 22.3 (3.2) | 22.1 (3.1) | 22.7 (3.1) | 23.1 (3.7) | 22.3 (3.5) | 23.1 (4.0) | 23.0 (3.8) |
| Baseline oropharyngeal eluate viral density | 5.4 (1.3) | 5.6 (1.2) | 5.6 (1.0) | 5.5 (1.1) | 5.5 (1.0) | 5.6 (1.3) | 5.6 (1.1) | 5.4 (1.2) |

| | | | | | | | | |
|---|---|---|---|---|---|---|---|---|
| ($log_{10}$ copies per mL) (SD) | | | | | | | | |
| Symptom onset (days) (SD) | 2.2 (0.8) | 2.3 (0.8) | 2.2 (0.8) | 2.4 (0.8) | 2.1 (0.7) | 2.2 (0.8) | 2.1 (0.7) | 1.9 (0.7) |
| Vaccinated (%) | 197 (99.5%) | 43 (97.7%) | 85 (96.6%) | 64 (95.5%) | 112 (98.2%) | 116 (100.0%) | 65 (98.5%) | 88 (97.8%) |
| **SARS CoV2 variants** | | | | | | | | |
| BA.1 (%) | 13 (6.6%) | 14 (31.8%) | 15 (17.0%) | 20 (29.9%) | 21 (18.4%) | 0 (0.0%) | 0 (0.0%) | 0 (0.0%) |
| BA.2 (%) | 52 (26.3%) | 18 (40.9%) | 30 (34.1%) | 37 (55.2%) | 42 (36.8%) | 24 (20.7%) | 5 (7.6%) | 1 (1.1%) |
| BA.2.3.20 (%) | 0 (0.0%) | 0 (0.0%) | 0 (0.0%) | 0 (0.0%) | 0 (0.0%) | 1 (0.9%) | 0 (0.0%) | 0 (0.0%) |
| BA.2.75 (%) | 41 (20.7%) | 0 (0.0%) | 5 (5.7%) | 0 (0.0%) | 5 (4.4%) | 34 (29.3%) | 27 (40.9%) | 30 (33.3%) |
| BA.4 (%) | 2 (1.0%) | 0 (0.0%) | 0 (0.0%) | 0 (0.0%) | 3 (2.6%) | 0 (0.0%) | 2 (3.0%) | 3 (3.3%) |
| BA.5 (%) | 42 (21.2%) | 0 (0.0%) | 25 (28.4%) | 0 (0.0%) | 32 (28.1%) | 31 (26.7%) | 28 (42.4%) | 26 (28.9%) |
| BN.1.9 (%) | 2 (1.0%) | 0 (0.0%) | 0 (0.0%) | 0 (0.0%) | 0 (0.0%) | 0 (0.0%) | 1 (1.5%) | 0 (0.0%) |
| Delta | 10 (5.1%) | 12 (27.3%) | 13 (14.8%) | 10 (14.9%) | 11 (9.6%) | 0 (0.0%) | 0 (0.0%) | 0 (0.0%) |
| Other(%) | 0 (0.0) | 0 (0.0%) | 0 (0.0%) | 0 (0.0%) | 0 (0.0%) | 2 (1.7) | 0 (0.0%) | 1 (1.1%) |
| XBB (%) | 10 (5.1%) | 0 (0.0%) | 0 (0.0%) | 0 (0.0%) | 0 (0.0%) | 9 (7.8%) | 3 (4.5%) | 3 (3.3%) |
| XBB.1.5-like (%) | 26 (13.1%) | 0 (0.0%) | 0 (0.0%) | 0 (0.0%) | 0 (0.0%) | 15 (12.9%) | 0 (0.0%) | 26 (28.9%) |

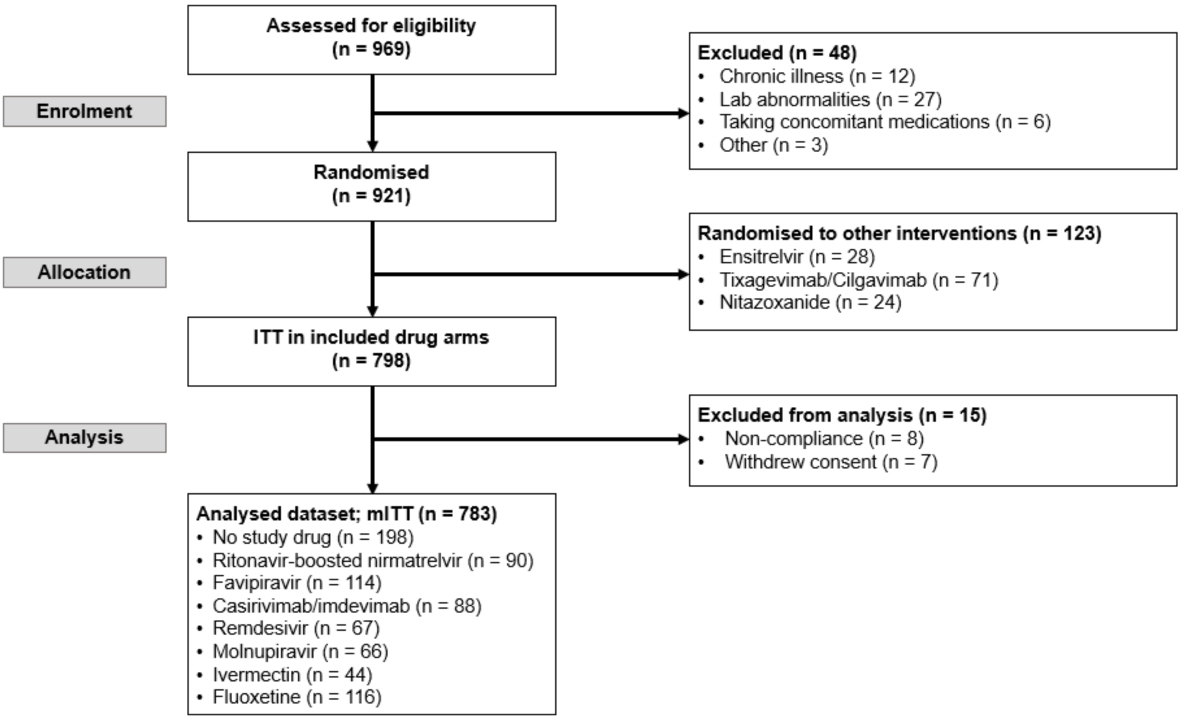

Figure S1: CONSORT diagram of meta-analysis of small molecule drugs and monoclonal antibodies

#### S12: Meta-analysis of fluoxetine and fluvoxamine

The clinical trial data examining whether SSRIs provide significant clinical benefit is mixed. A systematic review and post-hoc meta-analysis was performed of all outpatients randomised controlled trials evaluating fluoxetine or fluvoxamine. The inclusion criteria were – randomised controlled trials with patients with confirmed SARS-CoV-2, intervention in an outpatient population of fluoxetine of fluvoxamine, and an outcome including hospitalisation +/- death.

A search was made on 30.11.23 on PubMed and EMBASE using the search terms "fluoxetine", "fluvoxamine" and "COVID-19" with the search restricted to randomised controlled trials. Risk ratios for the efficacy endpoint were calculated for hospitalization +/- mortality. No fluoxetine trials fitted the inclusion criteria so only fluvoxamine was assessed. Primary endpoints and the dosing regimen used varied across the studies. Two of the trials used combination therapy along with fluvoxamine.

Meta-analytic risk ratios and forest plots were drawn and written using the R package meta version 6.5 in R version 4.3.2. A common effects model was used to estimate the meta-analytic ratios.

Treatment with fluvoxamine was associated with a slight reduction in hospitalisation +/- mortality in COVID-19 patients which did not reach significance at the 5% level (RR=0.80, 95% CI (0.62,1.01), p-value = 0.0641) (Figure S1).

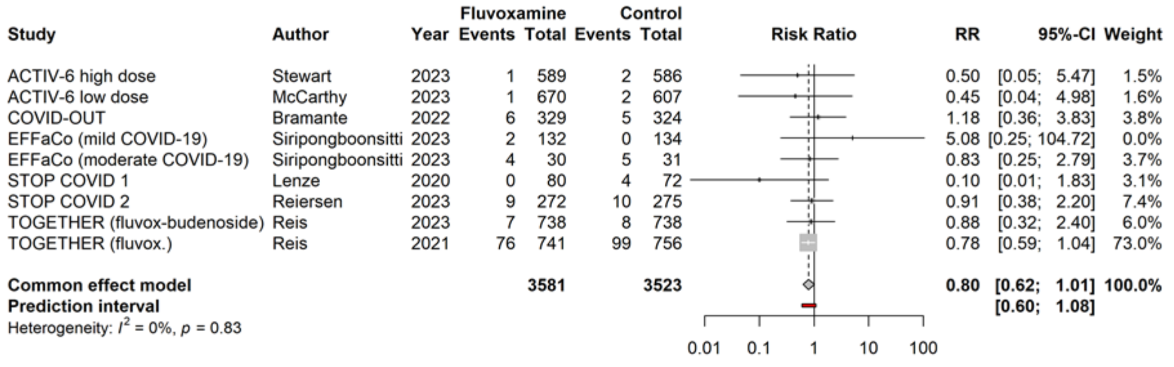

Figure S2: Forest plot of outpatient randomised controlled trials of fluvoxamine compared against standard of care, in mono or combination therapy.

### S13 Supplementary figures

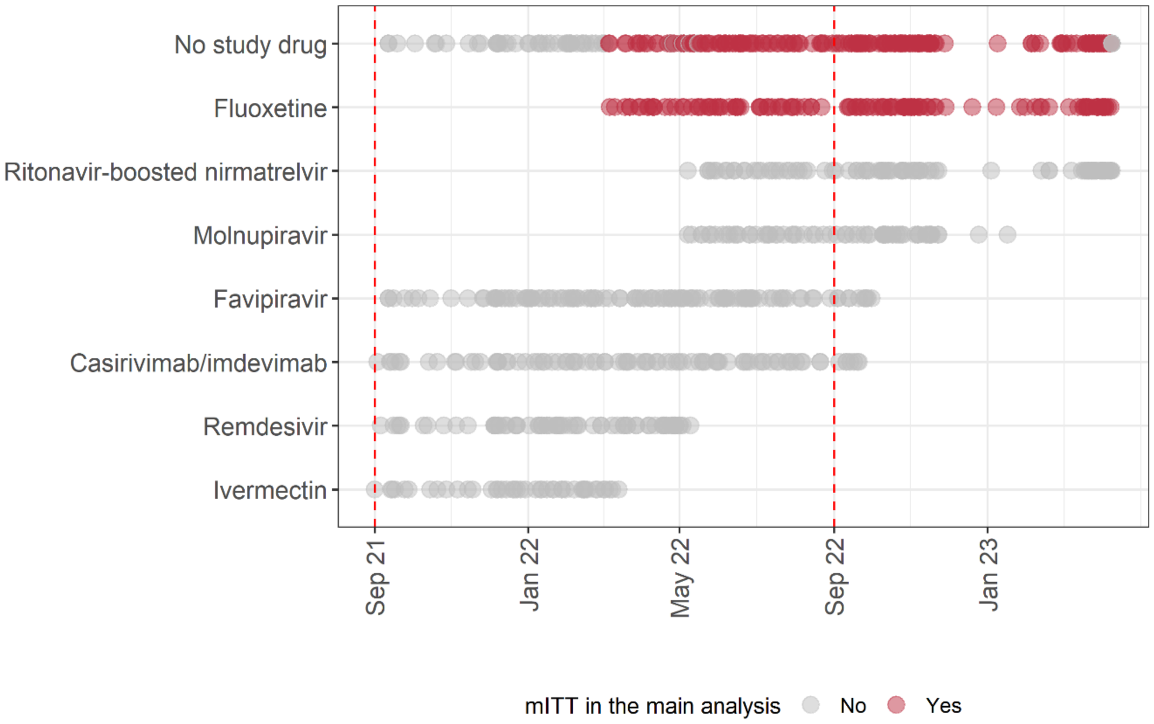

Figure S3: Randomisation date of 783 patients included in the meta-analysis. A total of 266 patients included in the main analysis (fluoxetine, and no-study-drug arms) were highlighted in red.

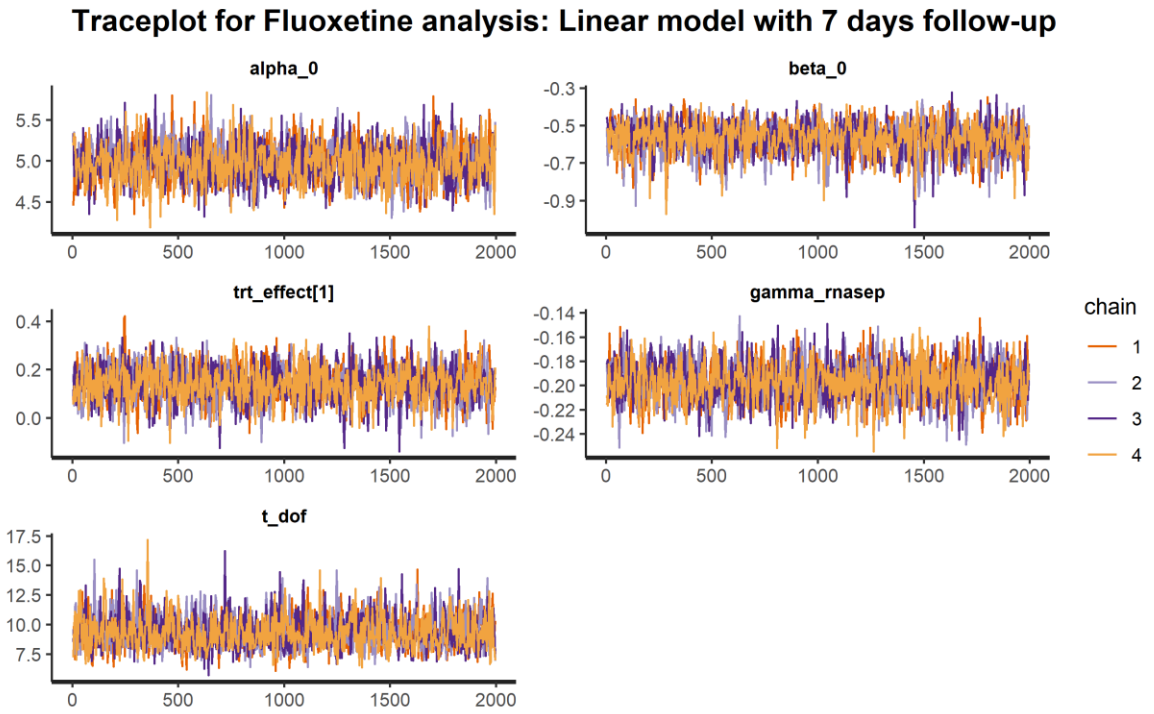

Figure S4: Traceplots depict convergence of four independent Markov Chain Monte Carlo chains for parameters in the main fluoxetine analysis with 7-day follow-up data.

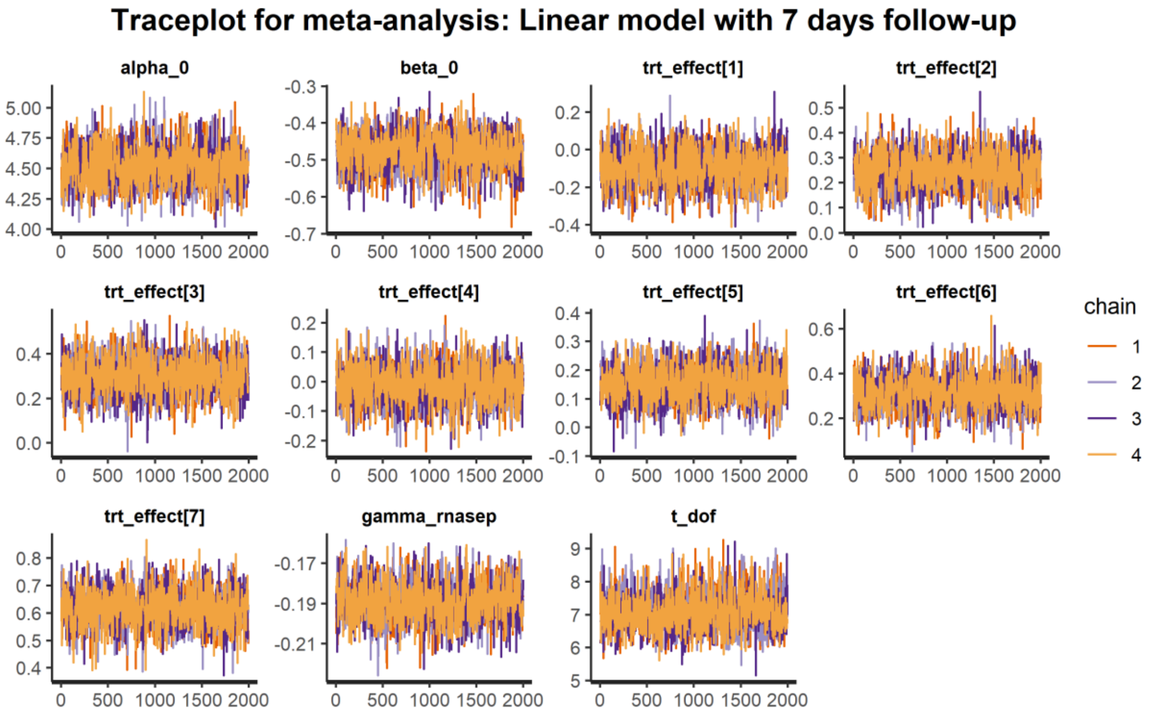

Figure S5: Traceplots depict convergence of four independent Markov Chain Monte Carlo chains for parameters in the meta-analysis with 7-day follow-up data.

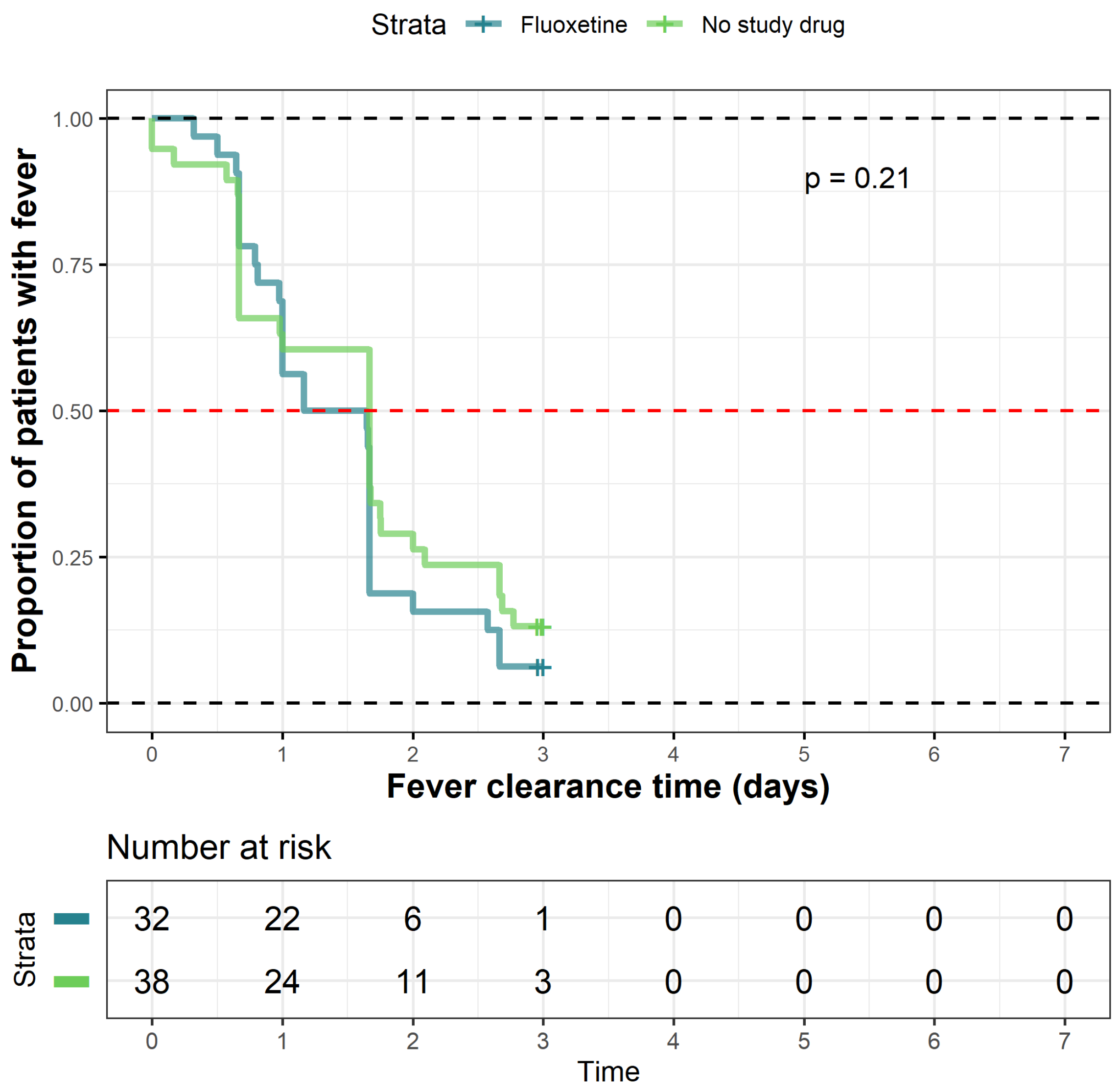

Figure S6: Fever clearance in patients who had a fever at baseline (>37.5 °C). All temperatures are axillary. Fever clearance is defined as time when temperature goes ≤ 37 °C for at least 24 hours. There was no significant difference in survival curves between the three arms (p=0.21). Plus signs indicate right-censored data.

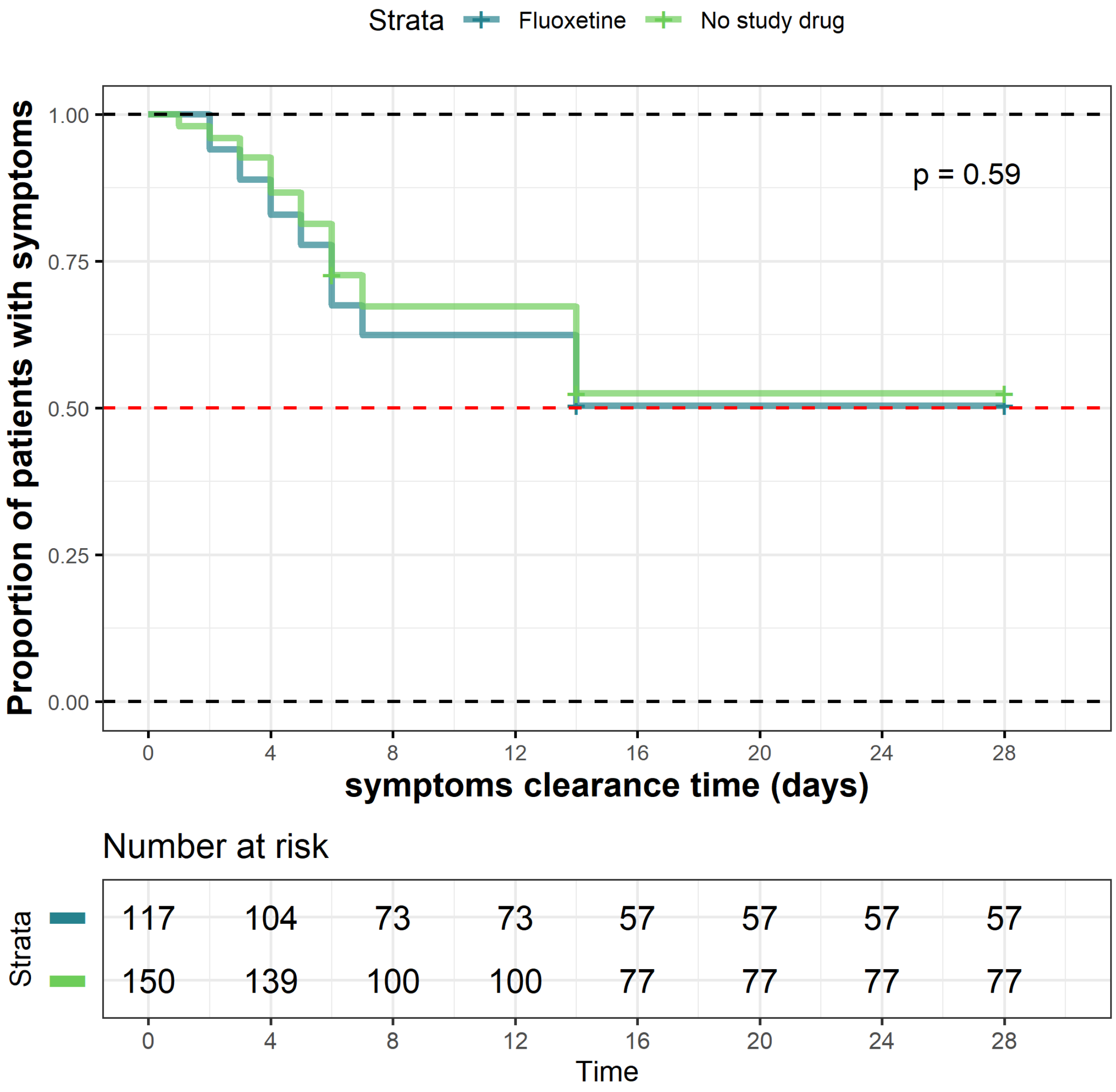

Figure S7: Symptom clearance in each arm in fluoxetine and no study drug analysis. Symptom resolution is defined as no reported symptoms (see list of symptoms in section S4 above). 138 (52%) patients were right censored at day 28, the last scheduled visit in the study.

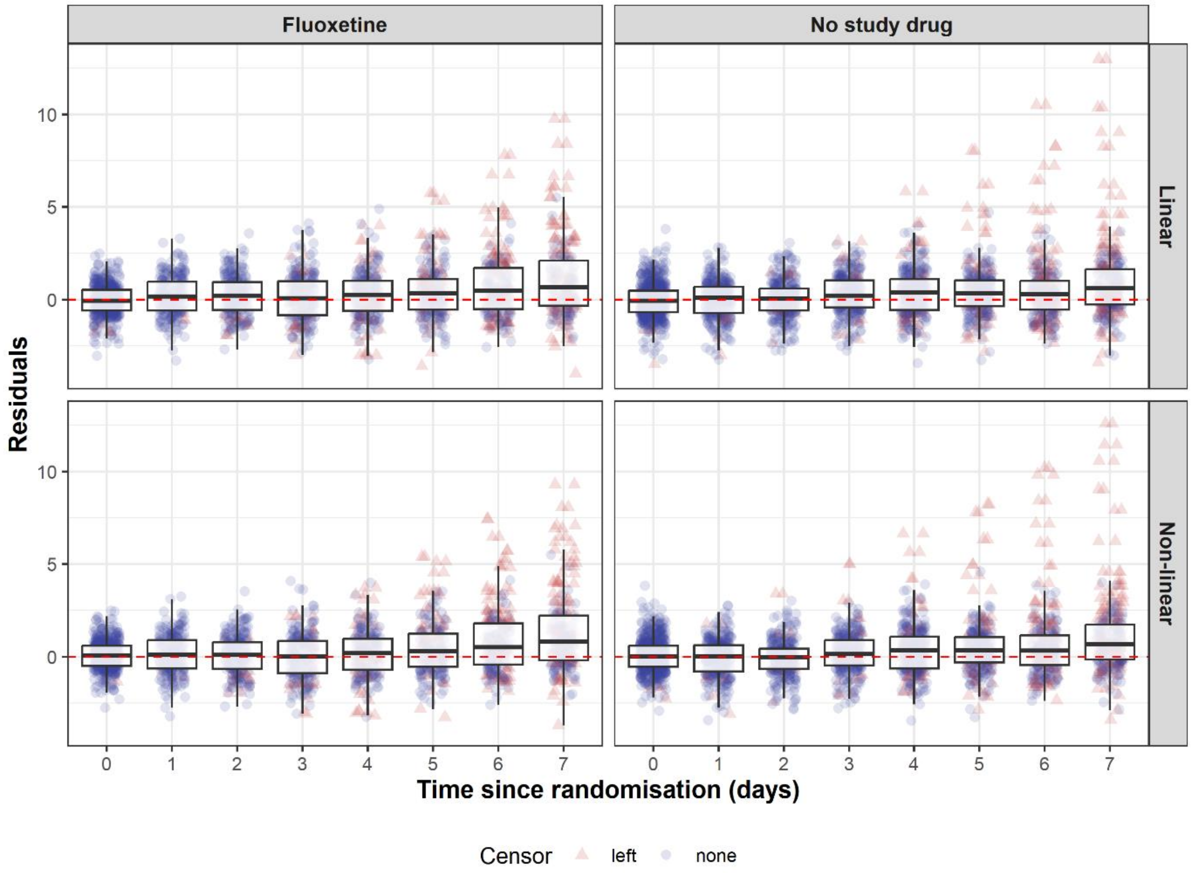

Figure S8: Analysis of the model residuals of linear model (the main model) and non-linear model as a function of time from randomisation in days. Residuals of left censored data points are shown as red triangles.

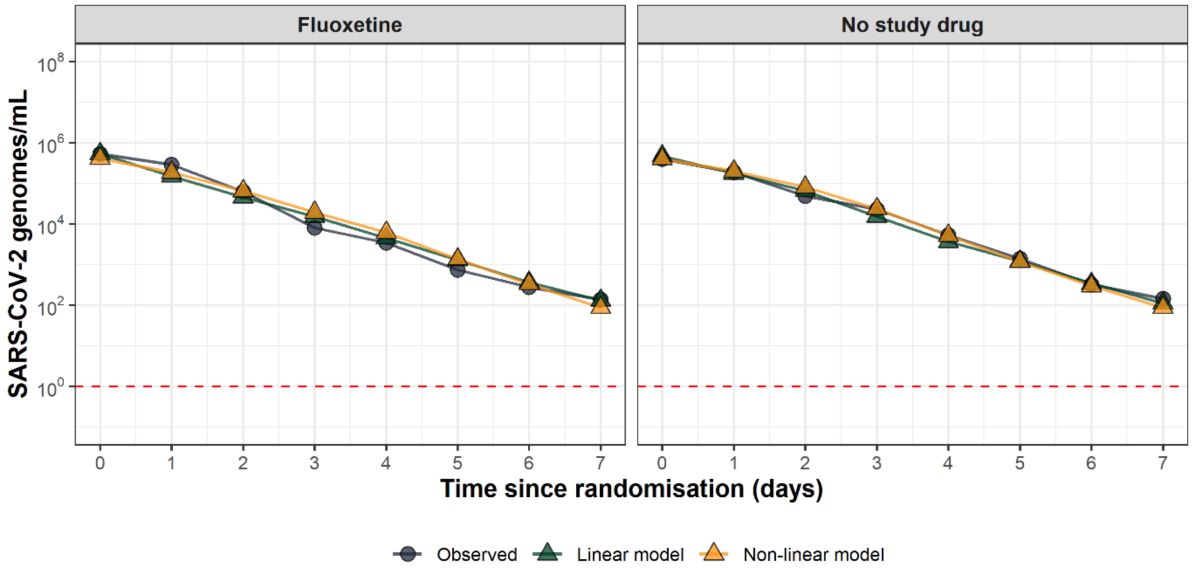

Figure S9: Daily median model predictions from the linear model (green triangles), non-linear model (yellow triangles) against the observed daily median values (black circles).
